# Electromyography recordings detect muscle activity before observable contractions in acute stroke care

**DOI:** 10.1101/2020.09.07.20190041

**Authors:** Christina Papazian, Nick A. Baicoianu, Keshia M. Peters, Heather Feldner, Katherine M. Steele

## Abstract

**Objective:** To determine whether electromyography (EMG) can be used in acute stroke care to identify muscle activity in patients with no observable activity during clinical examination.

**Design:** Stroke survivors admitted to a level one trauma hospital with initial NIH Stroke Scale scores of two or higher for arm function were recruited within five days of stroke (average 3±1 days), including eleven stroke survivors (7 male/4 female, age 56±11) with no observable or palpable arm muscle activity (Manual Muscle Test, MMT=0) and ten stroke survivors (6 male/4 female, age 64±1) with observable muscle activity (MMT>0). We placed wireless EMG sensors on five major muscle groups (anterior deltoid, biceps, triceps, wrist extensors, and wrist flexors) of the impaired arm for 3-4 hours during standard care.

**Results:** We were able to identify muscle contractions in all five muscles for all participants from EMG recordings. Contractions were easily identified from 30-minutes of monitoring for participants with MMT>0, but up to three hours of monitoring was required for participants with MMT=0 to detect contractions in all five muscles during standard care. Only the wrist extensors demonstrated significantly larger amplitude contractions for participants with MMT>0 than MMT=0. Co-contraction was rare, involving less than 10% of contractions. Co-contraction of two muscles most commonly aligned with the flexor synergy pattern commonly observed after stroke. For participants with MMT=0, number of contractions and maximum amplitude in acute care were moderately correlated with MMT scores at follow-up.

**Conclusion:** Muscle activity can be detected with surface EMG recordings during standard care, even for stroke survivors with no observable activity by clinical exam.

## Introduction

The initial days after stroke are a time of rapid change and uncertainty. While 80% of stroke survivors initially present with arm or hand impairments,^1^ early prognosis for motor recovery remains challenging.^2^, ^3^ Clinicians routinely assess these impairments by measuring an individual’s ability to volitionally move their body.^4^ Manual Muscle Testing (MMT) is recommended for initial assessments, since it takes less than five minutes and provides a standardized measure of motor function.^5^ However, for individuals with no volitional movement, the MMT alone cannot determine whether muscle activity is present. An MMT score of zero can indicate severe paresis with no muscle activity, that the muscle activity is below the level necessary to produce observable contractions, or that the patient was unable to fully participate in the assessment. For patients with MMT = 0, this can be frustrating as they take a “wait-and-see” approach to determine whether muscle activity will return.^6^

Patterns of recovery can be highly variable and unpredictable during the first week after stroke.^7^ Much of our current knowledge of early recovery is still based upon Thomas Twitchell’s observations from the 1950s using surface electromyography (EMG) recordings to monitor muscle activity.^8^ He used EMG recordings from major arm muscles to outline common patterns of recovery. He documented that flexion was often first observed and recovery generally progressed from proximal to distal joints, although patterns of recovery were highly variable. Of the 25 patients he followed, 17 could not initially move their arm. In these patients, he often could not detect muscle activity with EMG, but only monitored for short periods during specific movements. Whether modern EMG sensors or extended monitoring could identify contractions for these patients remains unknown.

Despite significant technical advancements, there are still very few studies of motor function during the initial weeks after stroke,^9^–^11^ with most relying on clinical tests like the MMT or observation. The presence of shoulder abduction and finger extension at 72 hours after stroke have been suggested as indicators of good recovery potential, whereas dense hemiplegia is thought to be an indicator of poor long-term recovery.^12^, ^13^ However, Prager and Lang (2012) found that initial paresis at day three only accounts for 28% of upper extremity function at three months post stroke.^14^ Stroke survivors are typically discharged from acute care within seven days after stroke, which requires clinicians to make decisions about discharge destinations, therapy interventions, and prognosis based on these early assessments.^5^, ^15^ This makes the need for more detailed and informative assessments during acute care a priority to further our understanding of motor recovery and prognosis.

Wearable sensors may provide more detailed quantitative measurements in acute care.^16^ Clinicians have recognized the potential of EMG as a prognostic tool, but challenges in evaluating EMG signals and deploying this technology have limited broader use.^17^, ^18^ In research, EMG has been shown to be superior to clinical assessments because it can identify changes in patterns of motor function that are not otherwise evident.^19^ Other wearable sensors, like accelerometers have been used extensively to monitor arm movement after stroke, including during inpatient rehabilitation and in the community.^20^, ^21^ While these sensors are useful for monitoring movement, they have limited use before muscle activity can produce movement and provide limited insight into muscle recruitment and coordination. EMG sensors have been developed that can be worn for extended periods and are integrated with sensors to simultaneously monitor movement.^22^–^25^ These technologies provide promising opportunities to expand upon Twitchell’s observations and use wearable sensors to enhance care.

The goal of this study was to investigate whether EMG can be used in acute stroke care to evaluate muscle activity, especially among patients with no observable activity or movement.

## Methods

We recruited 21 adult stroke survivors who had an upper-extremity motor impairment (score of two or higher on the NIH Stroke Scale for Motor Arm) within the first five days after admission to a level-one trauma hospital. This included 11 stroke survivors who had an MMT score of zero and 10 stroke survivors with upper-extremity impairment but MMT scores greater than zero (Table 1). MMT scores range from 0 (no observable or palpable contraction) to 5 (complete range of motion against gravity with full resistance). The participants with no observable muscle activity were 32-67 years old and included six who had experienced an ischemic stroke and five who had experienced an intraparenchymal hemorrhagic stroke. The participants with an MMT score greater than one were 29-85 years old and included eight who had experienced an ischemic stroke and two who had experienced an intraparenchymal hemorrhagic stroke. We obtained approval of the described protocols from the Institutional Review Board and all participants or their legal authorized representative provided informed consent.

Muscle activity and movement were monitored with wireless sensors placed on the impaired arm. We used the BioStamp RC sensors (BioStampRC, MC10, Lexington, MA), which allowed us to simultaneously monitor muscle activity from EMG recordings and movement from accelerometers. The Bluetooth interface and low profile (measuring 3.4 cm × 6.6 cm × 0.3 cm) also made these sensors appealing for extended monitoring without interfering with care. We placed the sensors on the anterior deltoid, biceps, triceps, wrist flexors, and wrist extensors of the impaired upper extremity using SENIAM guidelines and palpation (Figure 1). The sensor area on the upper extremity was shaved, if necessary. The sensors were secured to the skin with manufacturer-provided double-sided tape, and a strip of Tegaderm^TM^ transparent dressing was placed over each sensor in addition to Coban^TM^ self-adherent wrap around the arm to ensure sensors would not fall off or move during clinical care.

**Figure 1.**
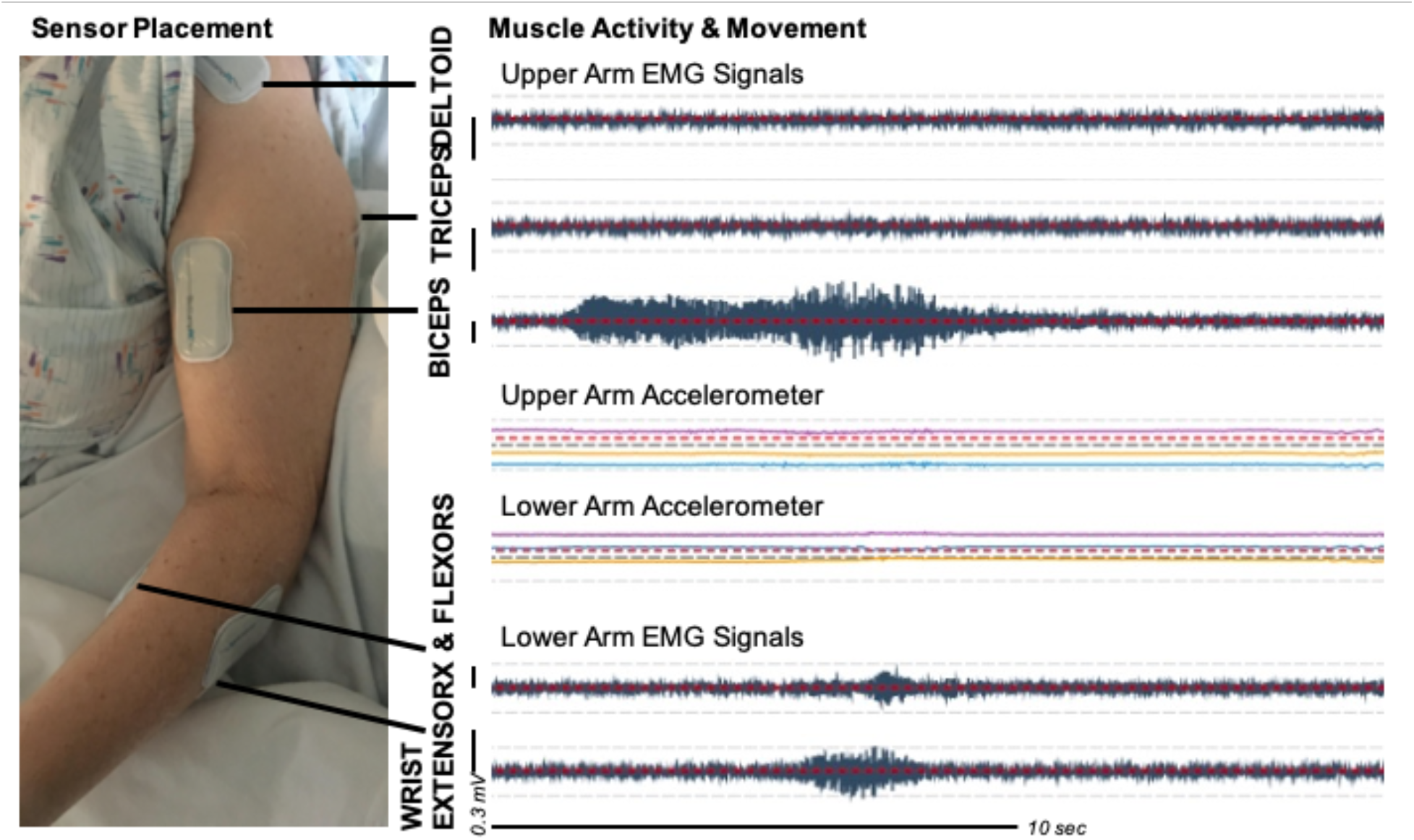
The Biostamp RC sensors were placed on five muscles – the anterior deltoid, biceps, triceps, wrist flexors, and wrist extensors – of the affected arm. Each sensor monitored muscle activity with EMG recordings and movement with a tri-axial accelerometer. An example contraction is shown for one of the MMT = 0 participants while they were watching TV in their hospital room. Accelerometer recordings from the upper arm and forearm show no movement, while the biceps had an extended contraction with some wrist flexor and extensor activity.

The sensors were left on for up to four hours, during which the participant continued with standard care. An activity check was performed every 30 minutes to briefly document activity (*e.g*., watching TV). For participants with MMT = 0, at the start of collection, we also performed an MMT exam to confirm the clinician’s documented assessment. The researcher asked the participant to attempt to move their arm. If no movement or contraction by palpation was detected, the researcher then performed a passive range of motion exam, repeatedly moving the shoulder, elbow, and wrist sequentially through their range of motion three times.

Raw EMG data were recorded at 1000 Hz. EMG data were processed using a fourth-order Butterworth bandpass filter between 20 and 400 Hz. Outlier data were discarded to remove high-magnitude hardware artifacts. Rectified EMG data were then smoothed using a moving median. The smoothed signal was used to manually identify contractions. For each participant, the EMG and accelerometer recordings from all muscles were evaluated concurrently in 5-10 second windows and the start and end time of each contraction was documented when the EMG signal’s magnitude exceeded baseline noise. The concurrent raw accelerometer data was also used to classify whether movement of the arm was present during each contraction. For all participants, contractions were identified during a 30-minute time period during standard care when the participant was awake and not involved in therapy. Additional 30-minute time periods were evaluated until contractions were identified from all five muscles or no data remained for analysis.

For each contraction, we calculated the maximum amplitude, median amplitude, and duration from the manually identified start and end times. We compared the contractions identified with and without movement for each group using the non-parametric Wilcoxon Signed-Rank Test. We compared contractions between participants with MMT = 0 and MMT>0 using the non-parametric Wilcoxon Rank Sum Test. We also evaluated co-contraction by evaluating whether multiple muscles were active at the same time. For five muscles, there are 31 potential co-contraction combinations: individual activation of each muscle, ten pairs of muscles, ten sets of three muscles, five sets of four muscles, or all five muscles. All analyses were conducted in Matlab (MathWorks Inc., Natick, Massachusetts, United States).

## Results

We were able to identify contractions from EMG recordings for all five muscles in all participants during standard care (Figure 2). For the participants with observable muscle activity (MMT>0), contractions were identified from all muscles in just 30 minutes of monitoring while awake in their hospital room. For five of the participants with no observable muscle activity (MMT = 0), we were also able to identify contractions from a single 30-minute time period. The other participants with MMT = 0 (N = 6) required monitoring for an additional 30 to 150 minutes before contractions were identified in all five muscles, with the biceps and triceps being the most difficult to identify contractions.

**Figure 2.**
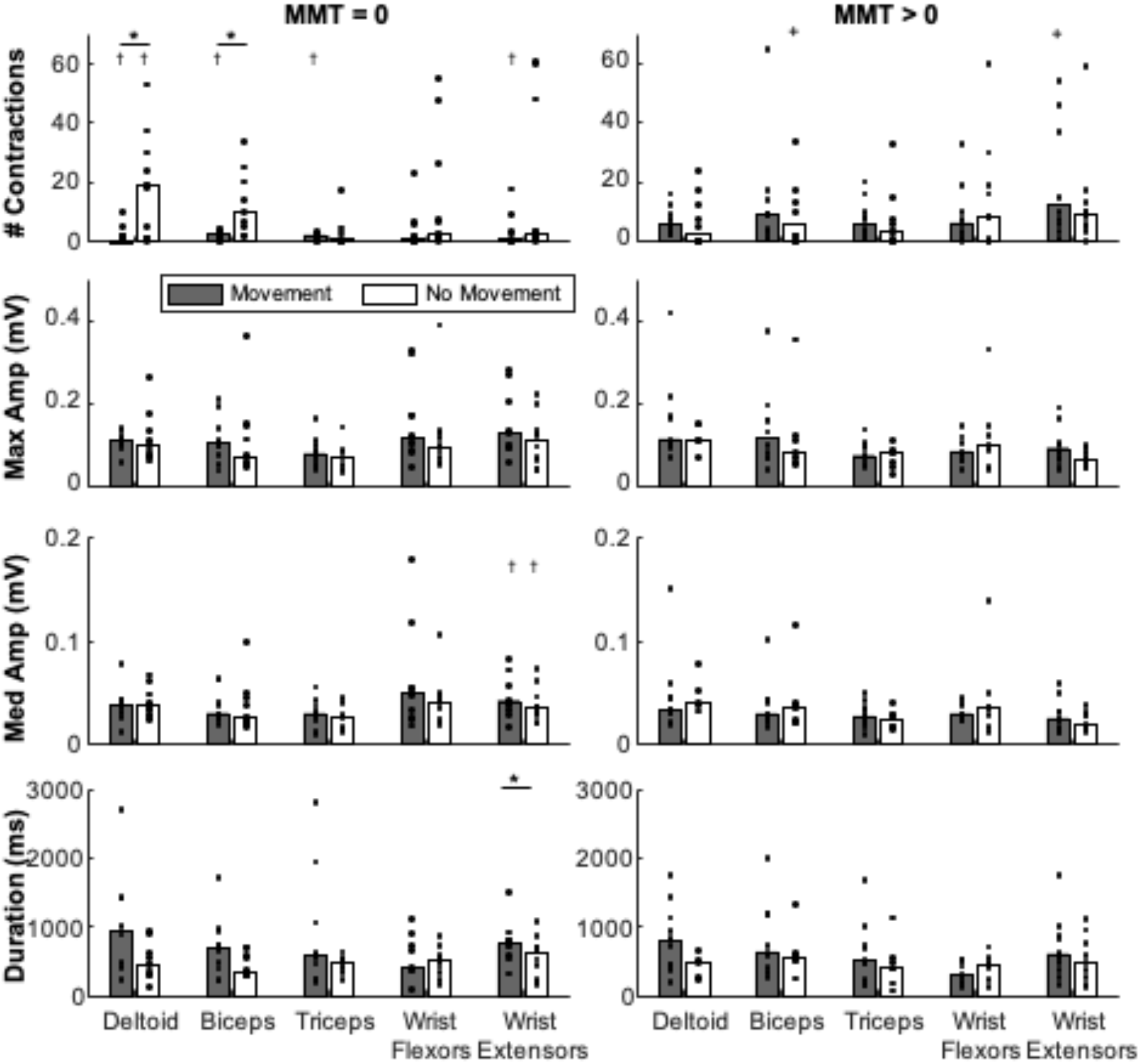
Contractions identified from EMG recordings for participants with no observable muscle activity (MMT = 0) and participants with some arm movement (MMT>0). The number of contractions per 30-minute time window, maximum amplitude, median amplitude, and contraction duration are shown for contractions with or without movement. Bars represent the median value, with black dots showing individual participants. All participants had contractions in all five muscles, although they not all participants had contractions both with and without movement. + Two participants with MMT>0 exceeded graph limits for number of contractions: one had 137 biceps contractions without movement and one had 108 wrist extensor contractions with movement. * Significant difference between contractions with or without movement. † Significant difference between participants with MMT = 0 and MMT>0.

The accelerometer on the sensors could also be used to detect whether contractions occurred with or without movement. As would be expected, the large majority of contractions for the participants with no observable muscle activity (MMT = 0) were during times of no movement. Contractions during times of movement among these participants likely reflect events when someone else was moving their arm (*e.g*., nurse repositioning in bed). This movement may have triggered reflex activity or some voluntary activity when the patient was trying to assist. Participants with observable muscle activity (MMT>0) had significantly more contractions with movement for all muscles except the wrist flexors (p = 0.003-0.01). For these participants (MMT>0), the majority of contractions occurred with movement (59-66% of contractions, except 45% for the wrist flexors) and higher MMT scores were associated with more contractions with movement (r^2^ = 0.16-0.42).

The maximum and median amplitudes of contractions were similar across muscles and between participants with and without observable muscle activity, with average maximum amplitude between 0.03 and 0.42 mV and median amplitude between 0.01 and 0.18 mV (Figure 2). Only the wrist extensors demonstrated significantly larger median amplitudes (p = 0.02) for participants with MMT>0 than those with MMT = 0. There were no significant differences in maximum (p = 0.08-0.99) or median (p = 0.16-0.81) amplitudes between contractions with or without movement. Durations of contractions were also not significantly different between groups, with most contractions lasting less than one second.

All participants were analyzed between one to five days after stroke. For the participants with MMT = 0, the number of contractions, maximum amplitude, and duration of contractions decreased with the number of days after stroke when the EMG recordings were made for all muscles except the wrist extensors (Figure 3). The association between days after stroke and contraction characteristics was much weaker for the participants with MMT>0. We also evaluated the correlation between the number of contractions, amplitude, and duration. For the participants with MMT = 0, individuals with more contractions had greater maximum amplitude (r^2^ = 0.16 – 0.62) and longer duration (r^2^ = 0.16 – 0.83) contractions. We observed weak or no correlation between the number of contractions, amplitude, and duration for participants with observable muscle activity (MMT>0), likely reflecting greater variety in activities that required different types of contractions.

**Figure 3.**
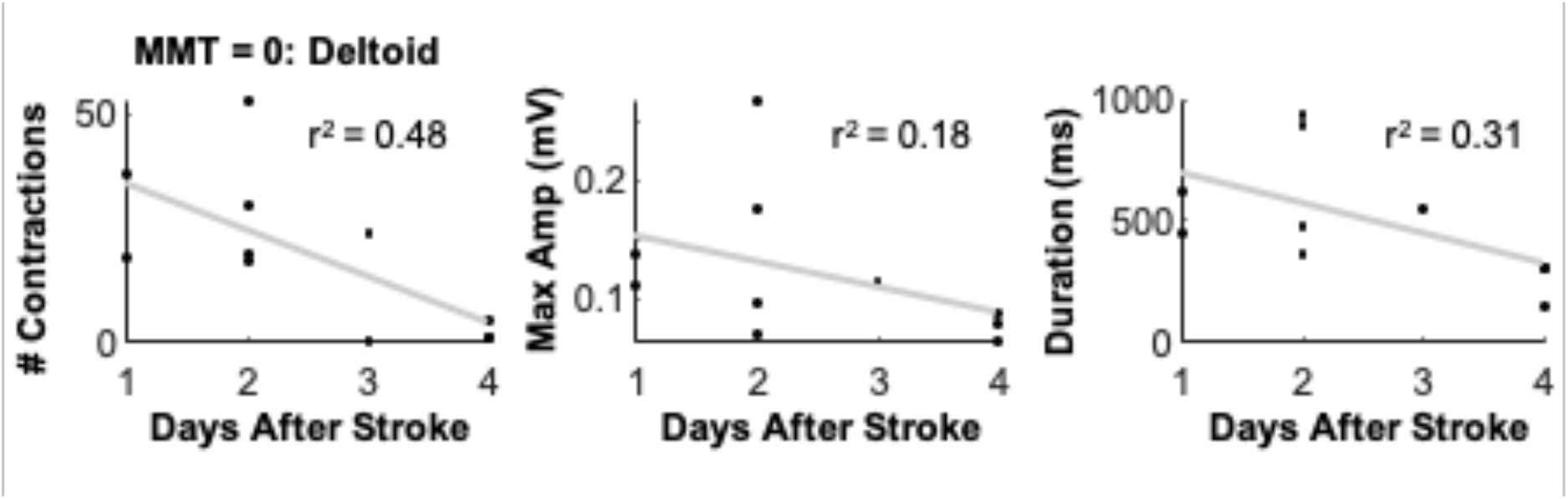
Contraction characteristics were dependent on the number of days after stroke that the EMG recordings were made for participants with MMT = 0. Data for the ten MMT = 0 participants who had deltoid contractions without movement are shown for the deltoid muscle. For all muscles except the wrist extensors, the number of contractions, maximum amplitude, and duration decreased with the number of days after stroke. For the wrist extensors, maximum amplitude increased with days after stroke and there were no associations between number of contractions or duration with days after stroke.

The participants with MMT = 0 were also monitored during the MMT and passive range of motion assessments (N = 9, Figure 4). While the MMT exam confirmed no muscle activity via observed movement or palpation, we identified contractions with EMG for all participants. At the beginning of the exam, when asked if they could move their arm, we identified muscle contractions among four of the participants, even though the examiner observed no movement or muscle activity. During the passive range of motion tests, muscle contractions were identified for all nine participants during elbow range of motion, and seven participants during both shoulder and wrist range of motion exams. As would be expected from reflexes, the deltoid was only active during attempted movement or shoulder range of motion.

**Figure 4.**
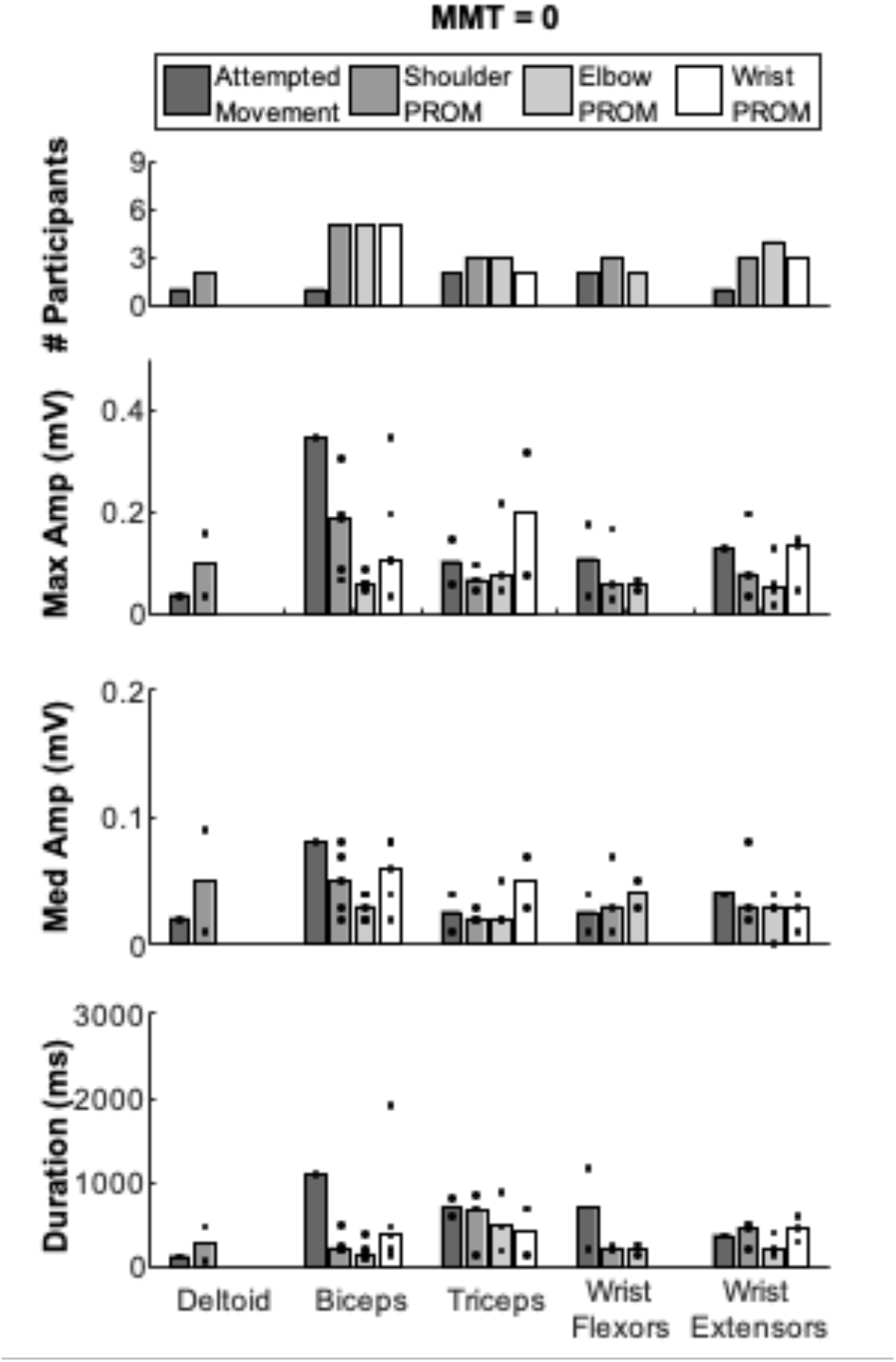
Contractions during the MMT and passive range of motion (PROM) assessments for the participants with MMT = 0. EMG recordings were collected for nine participants during these assessments. The number of participants who had identifiable contractions, as well as the maximum amplitude, median amplitude, and duration of contractions are shown for each muscle. Bars represent the median amplitude and duration across participants, with the black dots showing the median for individual participants.

Co-contraction was rare for all participants. For the participants with MMT = 0, over 95% of contractions involved only a single muscle (Figure 5). Co-contraction was significantly more common during contractions with movement (p = 0.003), which may reflect reflex activity when the arm was moved. Three of the four most common co-contraction patterns observed among the participants with MMT = 0 aligned with a flexion synergy, involving activation of the deltoid and wrist flexors (17% of cocontractions), biceps and wrist flexors (16% of co-contractions), or deltoid and biceps (11% of cocontractions). There were no co-contractions aligned with the extension synergy among participants with MMT = 0. Co-contraction was significantly more common among the participants with MMT>0 than MMT = 0, but still present in only 8% of identified contractions. For participants with MMT>0, cocontraction was more common during contractions with movement (p = 0.048) and the most common co-contractions were activation of the biceps and wrist extensors (23% of co-contractions) and activation of the wrist flexors and extensors (17% of co-contractions).

**Figure 5.**
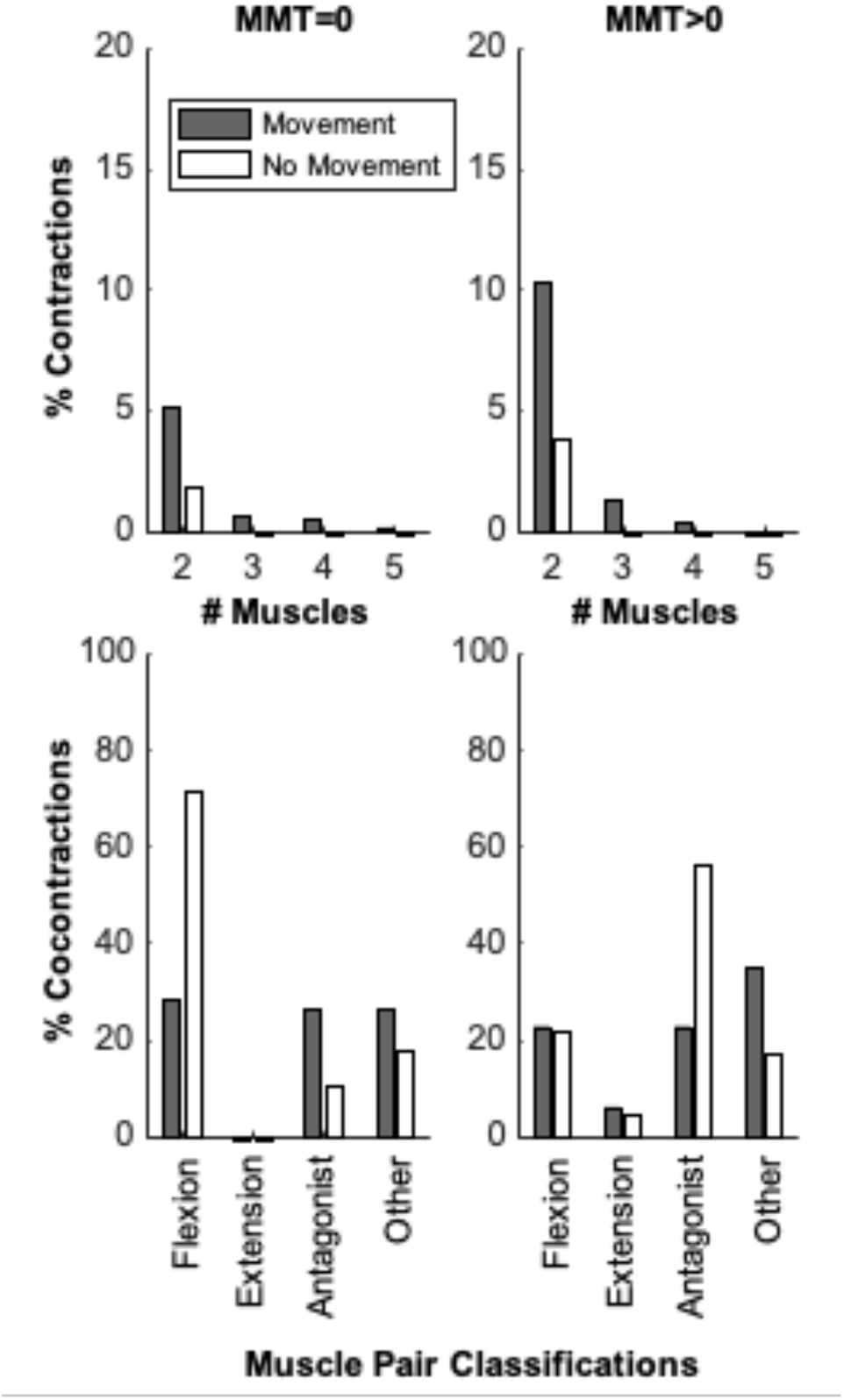
Co-contractions (percent of total number of contractions) were rare among participants without (MMT = 0) and with (MMT>0) observable muscle activity. For co-contraction involving two muscles, we classified them as aligning with the flexion or extensor synergy, antagonist pairs, or other patterns. The flexion synergy includes co-contraction of the deltoid, biceps, and wrist flexors. The extension synergy includes co-contraction of the triceps and wrist extensors.

All of the participants with MMT = 0 continued to inpatient rehabilitation (IPR) after acute care. At IPR discharge (range: 13-35 days after stroke), seven of the eleven participants still had an MMT score of one or less for all muscles. At follow-up (range: 1-12 months after stroke, average: 5 months), five participants had regained some arm function with MMT scores greater than two. The number of contractions and maximum amplitude at the initial evaluation were associated with MMT scores at follow-up for the biceps, triceps, and wrist flexors (Figure 6). Participants with fewer contractions and lower maximum amplitude in acute care had greater recovery at IPR follow-up. The wrist extensors had the opposite trends, where the few participants with more contractions, greater maximum amplitude, and longer duration contractions in acute care demonstrated the greatest improvements in MMT at follow-up.

**Figure 6.**
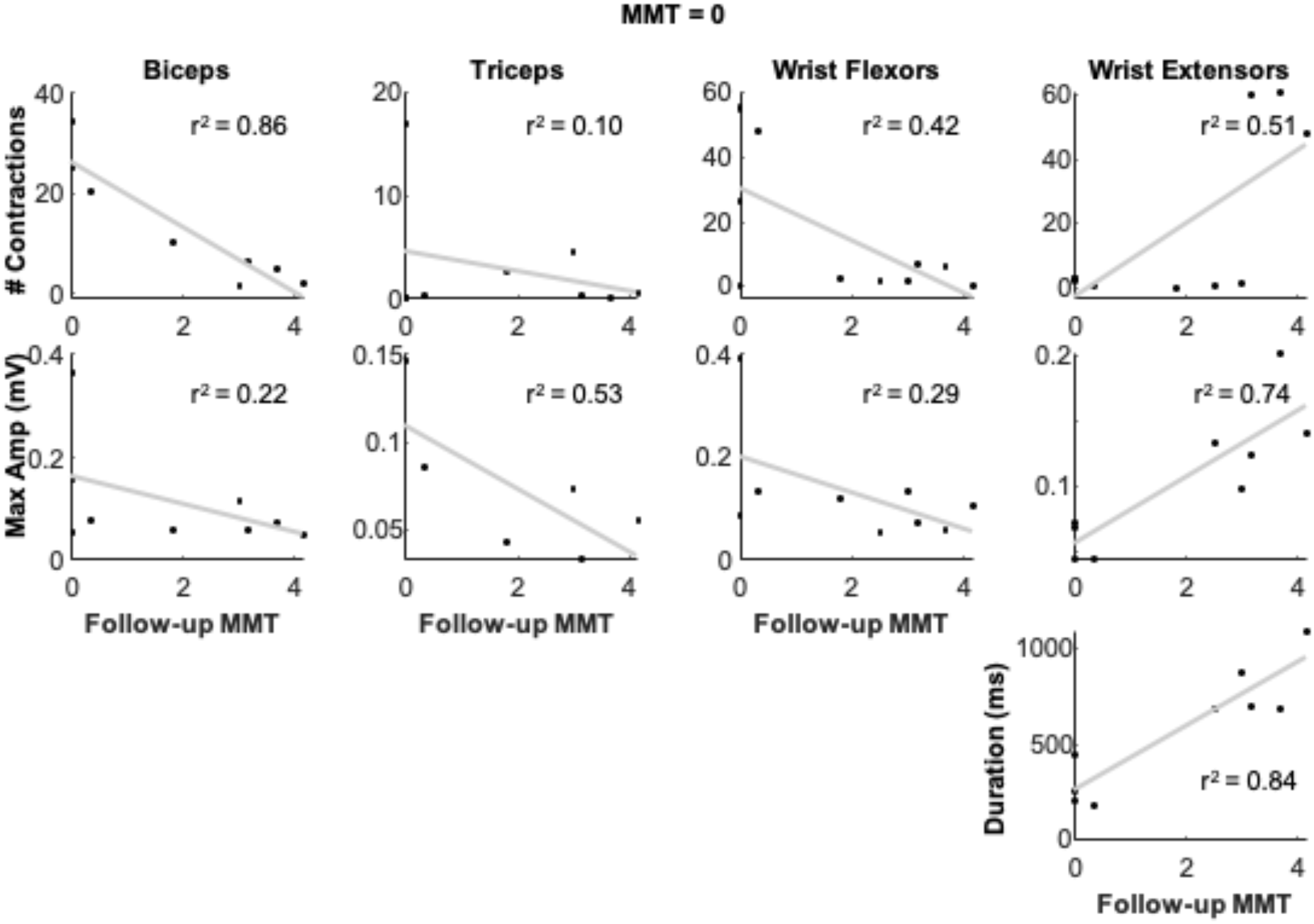
Contraction characteristics during contractions without movement were moderately associated with MMT scores at follow-up (1-12 months after stroke) among the participants with MMT = 0. For the biceps, triceps, and wrist flexors fewer contractions and lower maximum amplitudes in acute care were associated with greater improvement at follow-up. The wrist extensors had the opposite trend, where more contractions, higher amplitudes, and longer durations during acute care were associated with higher MMT scores at follow-up.

## Discussion

We found that monitoring muscle activity with EMG sensors during acute care could detect contractions in the major arm muscles of stroke survivors, even among those who had no observable activity from clinical exam. Surprisingly, we did not detect large differences in contraction characteristics – number of contractions, amplitude, or duration – between muscles or participants with varying arm function. This may reflect the wide variety of contractions that are captured when monitoring while a patient is in their hospital room, or that our signal quality was not good enough to detect more subtle differences. Regardless, the presence of contractions among all participants with MMT scores of zero indicates that EMG may help clinicians and patients evaluate muscle activity that is currently not observable with other methods to inform care and recovery.

The patterns of muscle activity we observed in acute care reflected those documented by Twitchell and others.^7^, ^26^–^28^ Like Twitchell’s observations,^8^ we saw wide variety in muscle activity after stroke, reflecting the unique deficits and recovery of each brain injury. Twitchell observed shoulder flexion emerging 6-38 days after stroke, with elbow and wrist flexion following shortly after. We also observed a large number of deltoid and biceps contractions in the first five days after stroke, which may precede the return of shoulder and elbow flexion. Patterns of co-contraction were also similar to previously documented patterns.^29^–^32^ The flexion synergy and antagonist co-contraction were the most prevalent co patterns. Participants with MMT = 0 demonstrated no contractions aligning with the extensor synergy, which is generally thought to appear later in recovery.

Adopting wearable sensors in acute care will ultimately require that these sensors provide unique and valuable insights that are not available with current methods.^33^ Detecting muscle activity alone may be sufficient – to address a patient’s question of whether or not their muscles are firing or to give therapists a tool to plan and evaluate their training sessions. For example, using EMG during the MMT or other clinical exams may help patients and clinicians see activity, encourage engagement, and give clinicians confidence in their assessment. Beyond detecting muscle activity, longitudinal evaluations will determine the diagnostic and prognostic value of EMG in acute care. Predicting future function has been attempted by many researchers.^34^–^37^ We found moderate correlations between contraction characteristics and future function for the participants with MMT = 0. Surprisingly for the biceps, triceps, and wrist flexors more and larger amplitude contractions were associated with worse outcomes at follow-up. These may reflect involuntary, reflex-driven contractions, but larger longitudinal studies will be required to link EMG-based measures with neurophysiology and recovery. Identifying opportunities for integrating EMG sensors into acute care can help support translation of this technology to the clinic.

## Study Limitations

For this research, we recruited a convenience sample with broad inclusion criteria to test the feasibility of deploying wearable sensors in acute stroke care. Evaluating specific types of stroke or groups with specific movement deficits may elucidate differences in contraction characteristics between individuals. We also relied on the MMT to evaluate arm function, which only provides a coarse ordinal scale that lacks the sensitivity to detect subtle improvements in strength.^38^–^40^ We chose to use this exam since it is conducted multiple times per day to evaluate function and inform clinical decision-making. Other exams, such as the Fugl-Meyer Assessment,^41^ could provide more detailed measures, but are not commonly used in acute care. To minimize disruption to care, we also chose to passively monitor with EMG, versus having the participant attempt to complete specific activities or observing their activities. More detailed records of the activities they were performing could further help decode and classify different types of contractions.

There were also numerous technical challenges that limited the methods we could use to process and evaluate the EMG data. The wearable sensors had an excellent form factor for use in the clinic, but had several limitations compared to research-grade systems that impacted signal quality.^42^ In particular, there was hardware interference that introduced noise, which made traditional processing techniques challenging. The inter-electrode distance was also wide, which increased crosstalk and limited our ability to target specific muscles.^43^ These factors strongly influenced our decision to manually identify contractions. While we attempted to automate classification using numerous prior methods,^44^–^48^ given the noise of the environment and technical limitations of the sensors, manual classification was the only method that provided sufficient confidence. We are confident that these limitations can be overcome, which will increase the potential value and impact of EMG in acute care.

## Conclusions

Muscle activity is present in the first week after stroke, even among participants with no signs of muscle activity during traditional exams. We were able to use EMG sensors to monitor and detect muscle activity for 21 stroke survivors with impaired arm function during acute care. Using EMG to detect contractions, evaluate common synergistic patterns, and track changes during routine care can provide new pathways to support recovery of stroke survivors.

## Data Availability

Please enquire with the corresponding author for data.

## Acknowledgements

The authors thank the clinical staff for their time and engagement with this research. We also thank Momona Yamagami and Ivana Milovanovic for their early support in launching this research. This research was supported by the National Institute of Biomedical Imaging and Bioengineering of the National Institutes of Health under award number R01EB021935.

